# Early diagnosis of transthyretin amyloidosis by detection of monomers in plasma microsamples using a protein crystal-based assay

**DOI:** 10.1101/2024.02.27.24303425

**Authors:** Diogo Costa-Rodrigues, José P. Leite, Maria João Saraiva, Maria Rosário Almeida, Luís Gales

## Abstract

Amyloid diseases are frequently associated with the appearance of an aberrant form of a protein, whose detection enables early diagnosis. In the case of transthyretin amyloidosis, the aberrant protein – the monomers – constitute the smallest species of the amyloid cascade, which creates engineering opportunities for sensing that remain virtually unexplored. Here, a two-step assay is devised, combining molecular sieving and immunodetection, for quantification of circulating monomeric transthyretin in the plasma. It is shown that mesoporous crystals built from biomolecules can selectively uptake transthyretin monomers up to measurable quantities. Furthermore, it was found that the use of endogenous molecules to produce the host framework drastically reduces unspecific adsorption of plasma proteins at the crystal surface, a feature that was observed with metal-organic frameworks. The assay was used to analyse plasma microsamples of patients and healthy controls. It shows a significant increase in the levels of monomeric transthyretin in the patients, proving its usefulness to establish the monomers as soluble and non-invasive marker of the disease. In addition, the assay can evaluate transthyretin stabilizers, an emergent strategy that broadened the treatment approach to the disease. Sensing the initial event of the transthyretin amyloid cascade with the proposed assay can make the difference for early diagnosis and eliminate the currently adopted invasive biopsies modalities for detection of the final products of the aggregation pathway.

## 1. Introduction

Transthyretin related amyloidosis (ATTR) is a highly debilitating and life-threatening disease. Significant variations in phenotypic and genotypic expression of the disease hamper early and accurate diagnosis which delays initiation of appropriate treatment (Adams et al. 2016). Current diagnostic methods include tissue biopsy analysis to detect amyloid deposits and genetic tests to search for TTR mutations. However, the disease can be associated with deposition of wild-type TTR in the myocardium which cannot be diagnosed by TTR sequencing.

Biopsy tests are focused on the observation of the final products of the amyloid cascade. Patients carry less stable TTR homotetramers that are prone to dissociation into non-native monomers, which in turn rapidly self-assemble into oligomers and, ultimately, amyloid fibrils. New methods for precise monitoring of the initial steps of the amyloid cascade, i.e. monomeric TTR levels in the plasma, are crucial to evaluate new pharmacological advances that broadened the treatment approach to ATTR, such as stabilizers of the native tetrameric TTR structure (Almeida et al. 2005) and antisense oligonucleotides that inhibit the hepatic production of TTR (Benson et al. 2018; Gales 2019).

Here, we devised a mesoporous crystal–based assay for selective extraction of TTR monomers over the native tetramers for posterior immunodetection (Fig. 1). The crystals work as sponges that enrich small proteins (including monomeric TTR) from a mixture, up to measurable quantities. By leveraging the molecular sieving properties of the mesoporous crystals and reducing unspecific surface adsorption of plasma proteins we developed an assay for accurate discrimination between the levels of circulating monomeric TTR in ATTR patients and healthy individuals.

**Fig. 1.**
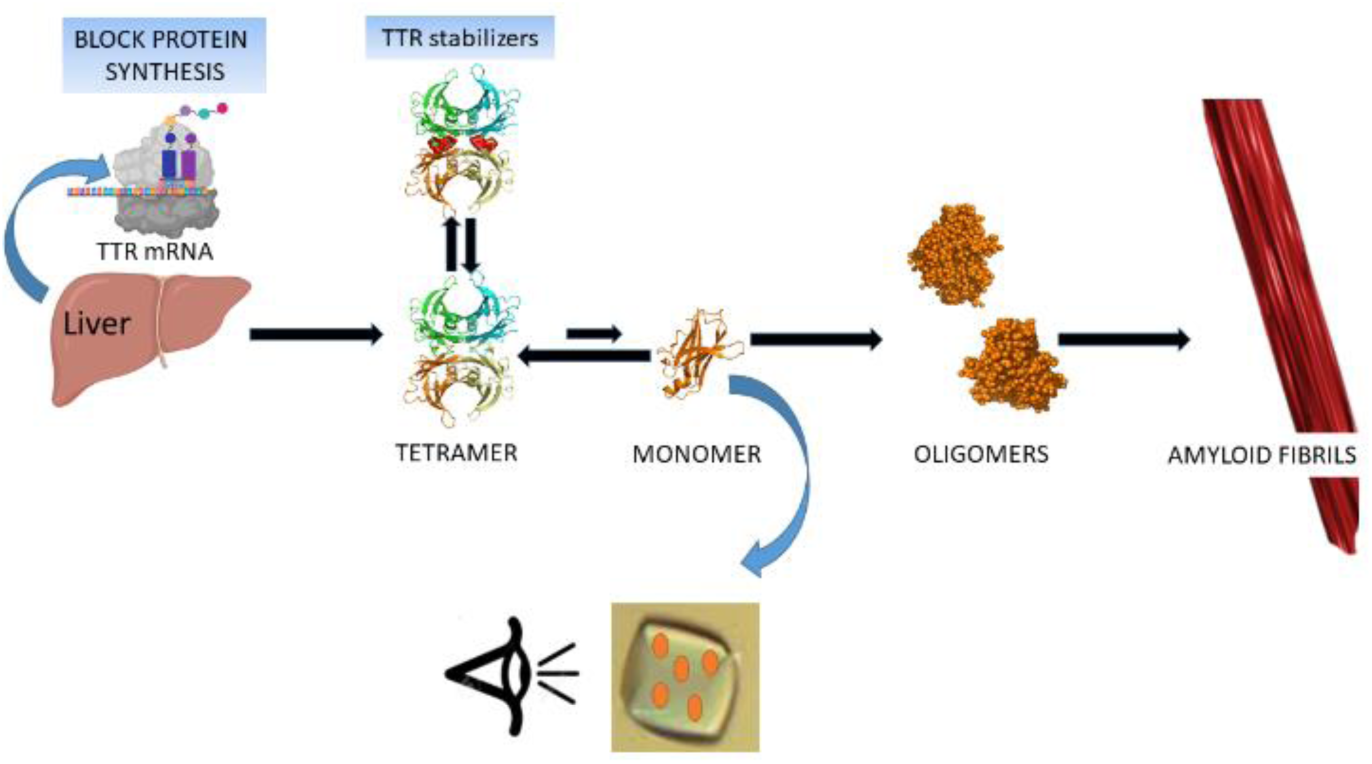
Schematic representation of the transthyretin amyloid cascade. Tetramer dissociation is the rate-limiting step. Emergent therapeutic strategies – inhibition of the synthesis of the protein and stabilization of the native protein – are pointed. Mesoporous materials will be used here for extracting the monomeric TTR species from plasma samples for subsequent immunosensing.

Crystals with high density of single sized pores are excellent candidates to separate mixtures of compounds with similar molecular sizes. Two different types of crystals were tested: protein crystals and Metal-Organic Frameworks (MOFs). A few researchers have been calling attention to protein crystals as microporous materials that offer a wide range of pore size (sometimes exceeding 10 nm), porosity (0.5-0.8), and pore surface area (800-2000 m^2^.g^−1^) (Vilenchik et al. 1998). Protein crystals are mostly produced to collect X-ray diffraction data to determine the atomic structure of the constituent molecules. Structural biologists are often insensible to the remarkable potential of such particles for engineering applications. Despite being soft and fragile solids that quickly dissolve in unfavourable environments, chemical cross-linking makes protein crystals much more stable, with little change in pore structure.

The most common strategies employed to adjust pore dimensions are size selection of the host protein and targeting specific network topologies. Several crystalline frameworks have been constructed with those strategies displaying a large range of pores, from subnanometer channels achieved by the assembly of short hydrophobic dipeptides (Görbitz 2001) up to proteins that self-assemble into 750 kDa cube-shaped cages with 13 nm inner cavities (Lai et al. 2014). The crystals have been used as scaffolds to investigate the diffusion in confined environments of small (Durão and Gales 2013) and large (Cvetkovic et al. 2005) guest molecules or as hosts to uptake compounds ranging from gas molecules (Afonso et al. 2010; Comotti et al. 2009) to biomacromolecules (Hashimoto et al. 2019). Among the biomolecules of interest to encapsulate are enzymes (Kowalski et al. 2019) whose stability is thereby enhanced. Another interesting field is the engineering of self-assembled porous protein crystals in living cells, aiming applications such as molecular recognition and storage of exogenous substances (Abe et al. 2017).

There are not many other crystalline materials that can reach sufficiently wide pores to encapsulate macromolecules. Another example is the class of MOFs. MOF materials are being developed for several biomedical applications, such as drug delivery, protection of biomolecules and cells, phototherapy and sensing (Mendes et al. 2020). In particular, MOF sensors perform at least as well as standard methods for many biomarkers (Dong et al. 2020; Gu et al. 2020; Sheta et al. 2019; Song et al. 2020; Wang et al. 2019; Wu et al. 2020; Zhou et al. 2018), including several amyloid related agents such as amyloid β peptide, α-synuclein, insulin, procalcitonin and prolactin (reviewed in (Leite et al. 2023; Tajahmadi et al. 2023)). These sensors reduce the overall complexity of currently used protocols, making them more appealing for a clinical setting. However, there is a focus in Alzheimer’s disease monitoring in detriment to other amyloidosis that are clearly under studied despite their relevance (e.g. Parkinson’s disease) or not explored at all (e.g. TTR amyloidosis) (Leite et al. 2023). Interestingly, in this work MOFs were not used as co-adjuvant materials as in most applications in construction of amyloid sensors but rather as a molecular sieve for extraction of the analyte.

## 2. Experimental Section

### 2.1. Materials

Thermolysin from *Geobacillus stearothermophilus* was purchased from Sigma-Aldrich (Type X, lyophilized powder, 30-350 units/mg protein (E1%/280)), resuspended as previously described (Hausrath and Matthews 2002) and used without further purification. Cytochrome c from bovine heart (≥95%) was purchase from Sigma-Aldrich. Terbium nitrate pentahydrate and 4,4′,4″-s-triazine-2,4,6-triyl-tribenzoic acid (H3TATB) were also purchased from Sigma-Aldrich (UK). All other reagents were of analytical grade.

### 2.2. Recombinant human TTR

TTR wild-type (TTR WT) and variants were produced using a bacterial expression system and purified as previously described (Furuya et al. 1991). Recombinant TTR was dialyzed against endotoxin-free phosphate-buffered saline (PBS), concentrated using Vivaspin ultrafiltration units (GE Healthcare, Chicago, IL, USA), and quantified using Bradford protein assay (Bio-Rad, Hercules, CA, USA).

### 2.3. Plasma Samples

The utilization of «human blood samples from individuals who requested to perform blood collection with predictive character at the Center for Predictive and Preventive Genetics (CGPP) of the IBMC to evaluate the condition of the pre-symptomatic V30M mutation» was submitted by the group Molecular Neurobiology (i3S) and approved by the Ethical Committee of the University of Porto (Report n°36/CEUP/2017).

### 2.4. Synthesis of Tb-MOF

Tb-MOF was synthesized as described in (Leite et al. 2019) which was adapted with minor modifications from (Park et al. 2007). Briefly, terbium nitrate (1.38 × 10^−4^ mol) and 4,4′,4″-s-triazine-2,4,6-triyl-tribenzoic acid (4.54 × 10^−4^mol) were dissolved a mixture of dimethylacetamide/methanol/water (4.0/0.8/0.2 mL, respectively). The solution was heated to 378 K for 48 h in a scintillation vial and then slowly cooled to ambient temperature. Colourless octahedral crystals were clearly visible.

### 2.5. Synthesis of cross-linked thermolysin crystals (TLN-CLC)

Tetragonal crystals were grown according to the method described in (Leite and Gales 2019), using the hanging drop vapour diffusion method in 24-well crystallization plates at 20°C. In each well, 500μl of 1M ammonium sulphate was added to the reservoir as precipitant solution. A drop of 6μl was added to the cover slide; its composition was 50% v/v of 150 mg·ml^−1^ thermolysin dissolved in 45% DMSO, and 50%v/v of 1M zinc chloride also dissolved in 45% DMSO. After ∼24h the presence of crystals was confirmed. Crystallization conditions were carefully controlled to obtain reproducible crystals. Crystal cross-linking: one drop of 1μl of 25% v/v glutaraldehyde was added directly to the crystal’s drop. The well was then closed again (well-sealed with grease to avoid vaporization of glutaraldehyde), and the crystals were left incubating ∼1h30min.

### 2.6. Analysis of the monomeric TTR extraction by the mesoporous crystals using SDS-PAGE

10 mg of MOF crystals were incubated overnight with recombinant TTR at 0.2 mg.ml^−1^ (150 μl sample volume) in PBS 1x, pH 7.4. Crystals were washed to remove superficially adsorbed protein, dissolved by gentle heating and acidification and resuspended in 50 mM HEPES pH 7.5. Samples were analysed in 15% (w/v) polyacrylamide gels by SDS-PAGE. NZYColour Protein Marker II (NZYtech) was used as a molecular weight marker. The same procedure was used with TLN-CLC but with a sample of five crystals.

### 2.7. Analysis of the monomeric TTR uptake by TLN-CLC using molecular probes cytochrome c and fluorescein

Single TLN-CLC crystals were incubated at RT, for 3 h, in 5 µl drops of TTR (3.6×10^−6^ M in phosphate buffered saline pH 7.4). The void volume of the crystals, which is expected to diminish by the uptake of TTR monomers, was estimated by two methods, schematically presented in Fig. 2.

**Fig. 2.**
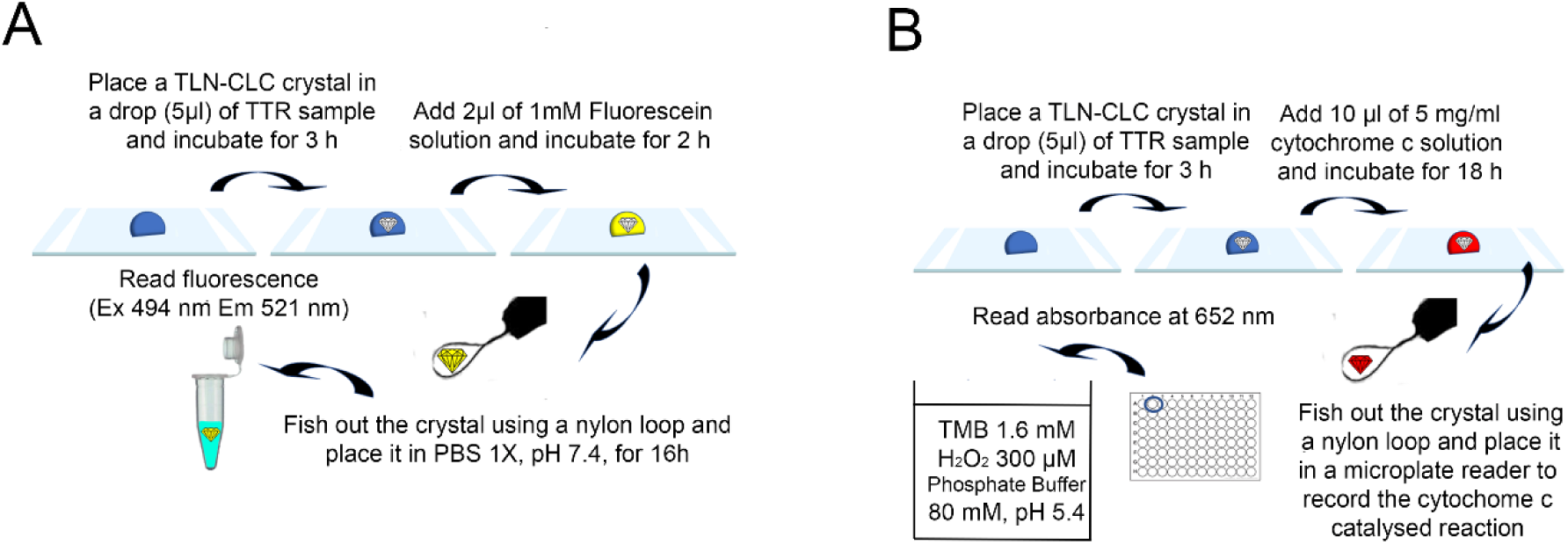
Schemes of the experimental procedure for estimation of the free void volume of the TLN-CLC crystals after TTR uptake, with fluorescein (A) and cytochrome c (B).

Method 1 (Fig. 2A): 2 µl of a solution of 1mM fluorescein was added to the crystal drop and incubated for 2 hours. The crystals were then fish out using a nylon loop, gently placed in a clean surface to remove the solution in the loop and transferred, also with a loop, to a phosphate buffered saline solution. Fluorescence was measured after 18h (Excitation at 494 nm and emission at 521 nm).

Method 2 (Fig. 2B): 10 µl of a solution of cytochrome c (5 mg.ml^−1^) was added to the crystal drop and incubated for 18h. The crystals were fished out with a nylon loop, quickly passed through a washing solution, and transferred to a reader plate to record a cytochrome c catalysed colorimetric reaction. Peroxidase activity was determined using 3,5′,5,5′-tetramethylbenzidine (TMB) as described in (Parakra et al. 2018). Briefly, cytochrome c samples in the presence of TMB (1.6 mM) were reacted with H_2_O_2_ (300 μM) in phosphate buffer (pH 5.4, 80 mM), at room temperature. TMB oxidation was follow measuring the absorbance at 652 nm and the initial velocity of the reaction determined.

Three independent experiments were performed for each condition. The morphology of TLN-CLC crystals was very homogeneous, which was confirmed by optical imaging of the crystals and by measuring their fluorescein adsorption capacity.

### 2.8. TLN-CLC based assay for evaluation of TTR drug candidates

Recombinant TTR samples were incubated overnight with tafamidis or acoramidis at molecular drug/TTR ratios of 10/1, 1/1 and 0/1. Then, the procedure described in section 2.7 was followed. Three independent experiments were performed for each condition.

### 2.9. TLN-CLC based assay for analysis of plasma samples

Three TLN-CLC crystals were incubated three hours at RT in 5 µl drops of plasma samples. The uptake of TTR monomers, was evaluated by immunoassay analysis, namely Western Blot and ELISA.

Western Blot: After incubation the crystals were fished out using a nylon loop, gently placed in a clean surface to remove the solution in the loop and transferred, to a PBS solution. SDS-PAGE analysis was performed (15% (w/v) polyacrylamide gels) and the proteins transferred onto nitrocellulose membranes. The membranes were blocked with 5% skim milk in PBS with Tween 20 (PBST), for 1 h at room temperature. Next, the membranes were incubated overnight at 4 °C, with a rabbit polyclonal antibody against human TTR (DAKO) at a 1:500 dilution. The membranes were washed three times in PBST for 5 min and were then incubated with a secondary antibody, conjugated with HRP (Goat anti-rabbit IgG, at a 1:10000 dilution) for 1 h at room temperature. Protein bands were detected via the enhanced chemiluminescence technique, using the Clarity ECL detection kit (BIO-RAD). Four independent experiments were performed for each condition.

ELISA: For determination of TTR inside the crystals, rabbit polyclonal anti-human TTR antibody (DAKO) was used as capture antibody at a 1:500 dilution, sheep anti-human TTR (Abcam) antibody diluted at 1:2500, as secondary antibody, and lastly was used donkey anti-sheep antibody (SIGMA) in a dilution of 3:10000. SIGMAFAST (p-nitrophenyl phosphate tablets) were used and absorbance was determined at 410 nm. Three independent experiments were performed for each condition.

### 2.10. Mass spectrometry (protein gel band, figure 3E)

Peptide cleavage characterization was performed by nanoLC–MS/MS, with a Ultimate 3000 liquid chromatography system coupled to a Q-Exactive Hybrid Quadrupole-Orbitrap mass spectrometer (Thermo Scientific, Bremen, Germany). Samples were loaded onto a trapping cartridge in a mobile phase of 2% ACN, 0.1% FA at 10 μL·min^−1^. After 3 min loading, the trap column was switched in-line to a reverse phase column at 300 nL·min^−1^. Separation was performed with a gradient of 0.1% FA (A) 80% ACN, 0.1% FA (B). The mass spectrometer was operated in a Full MS positive acquisition mode. The ESI spray voltage was 1.9 kV. The global settings were use lock masses best (m/z 445.12003), lock mass injection Full MS, chrom. peak width (FWHM) 30s. The full scan settings were 70k resolution (m/z 200), AGC target 3 × 106, maximum injection time 200 ms, scan range: 400–4000 m/z.

**Fig. 3.**
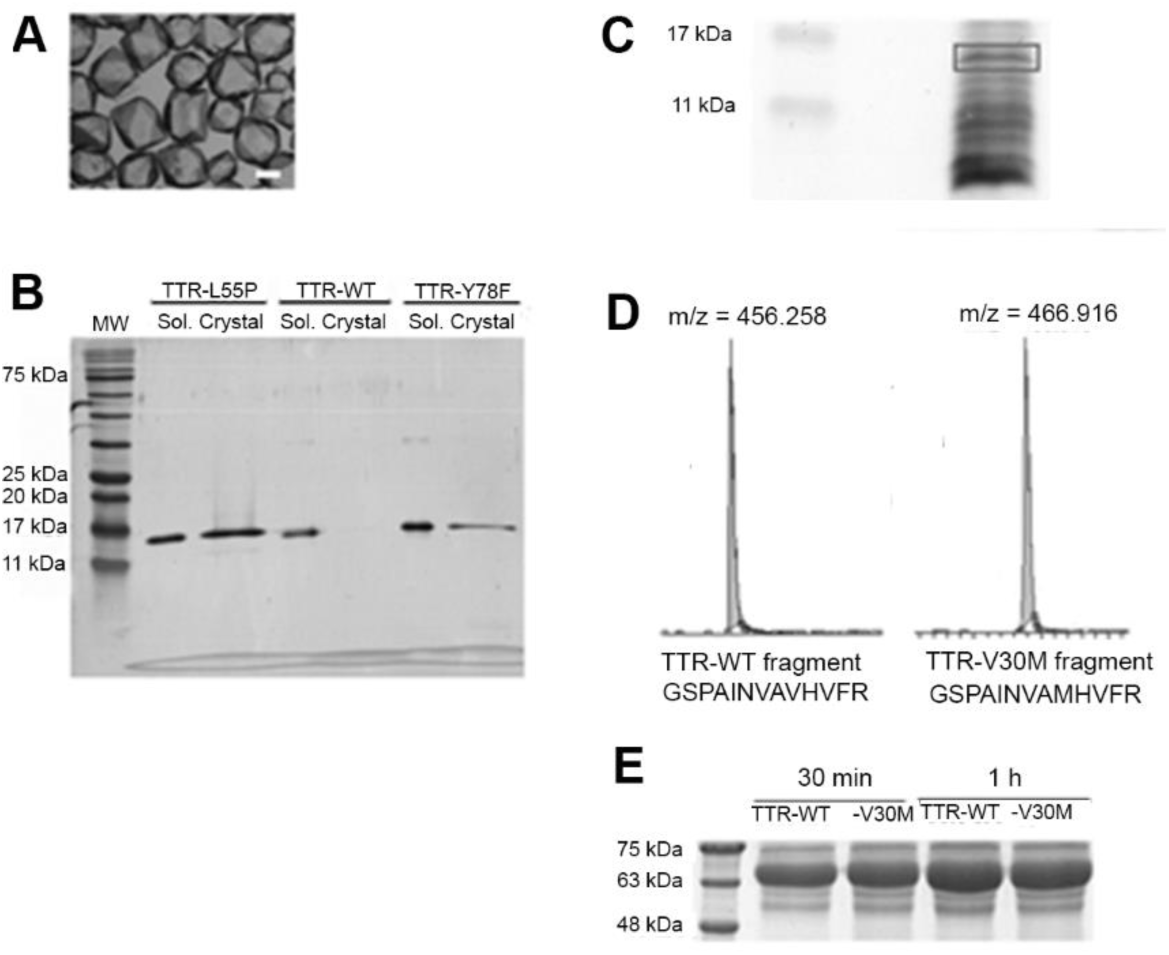
Analysis of the protein extracted from recombinant TTR samples and from plasma samples by Tb-MOF crystals. **A**. Optical image of Tb-MOF crystals (scale bar 0.5 mm). B. SDS analysis of the dissolved crystals after protein extraction from pure samples of recombinant TTR. Results shown for the TTR WT and for the amyloidogenic variants TTR-L55P and TTR-Y78F (Sol. - protein soaking solution and Crystal – protein extracted from the crystals dissolved after incubation with the soaking solutions). Tb-MOF extracted observable quantities of protein from the amyloidogenic variants, but not from the TTR WT (mostly tetrameric). **C**. SDS-PAGE gel of the protein extract from the plasma sample of a TTR-V30M carrier using Tb-MOF. The band ≈ 14kDa (approximate MW of the TTR monomer), highlighted in the figure, was excised and analysed by mass spectrometry, confirming it as TTR (**D**). **E**. SDS-PAGE gel of plasma proteins that become adsorbed at the external surface of Tb-MOF crystals, after 30 min and 1 h of incubation time with TTR-V30M and TTR-WT plasma samples. Size-excluded proteins become adsorbed which reduces the separation efficiency of the crystals.

### 2.11. Statistical analysis

Data was analysed using GraphPad Prism Software (version 9.5.0). Unpaired *t*-test with Welch’s correction was performed. Levels of statistical significance at * p < 0.05, ** p < 0.01, *** p < 0.001, **** p < 0.0001 were used. The number of independent experiments performed is given above, in the description of the respective assay.

## 3. Results and discussion

### 3.1. Tb-MOF separate TTR monomers from tetramers in vitro but are prone to unspecific surface adsorption of plasma proteins

Initial screening for MOFs capable of tailored uptake of TTR monomers and exclusion of tetramer led us to select Tb-MOF (Fig. 3A), a robust material, stable in several solvents, that has cages of 3.9 and 4.7 nm (Park et al. 2007). Screening consisted in the incubation of MOF crystals (10 mg) in samples of pure recombinant TTR variants (0.2 mg.ml^−1^, 150 μl), including the wild-type protein (TTR-WT) and two highly amyloidogenic variants: TTR-L55P (Bonifácio et al. 1996) and TTR-Y78F (Redondo et al. 2000). Crystals were then dissolved and analysed by SDS-PAGE. TTR was detected in all TTR soaking solutions, as expected (Fig. 3B). Crystals that were pre-incubated with the amyloidogenic variants TTR-L55P and TTR-Y78F encapsulated measurable quantities of monomeric TTR, while crystals soaked with TTR-WT did not. The amyloidogenic variants are less stable, more prone to dissociate into monomeric species that penetrate in the MOF framework.

The promising results obtained with pure recombinant TTR solutions prompted us to investigate the detection of TTR monomers in plasma samples of ATTR patients carrying TTR-V30M, the most frequent TTR variant associated with amyloidosis worldwide, found endemically in the northern parts of Sweden, and in regions of Portugal and Japan. We observed that a mixture of low molecular weight plasma proteins penetrates in the Tb-MOF framework, which was further separated by gel electrophoresis (Fig. 3C). The gel band with the expected molecular weight of the TTR monomer was excised and analyzed by mass spectrometry. TTR-V30M patients are heterozygotes and, accordingly, both the TTR-WT and the TTR-V30M monomer were identified (Fig. 3D).

Unfortunately, inspection of the crystals in contact with plasma samples with a stereomicroscope reveals the formation of a layer at the surface of the crystals composed by plasma proteins which may include tetrameric TTR. This external protein layer is hard to separate from the smaller protein species that occupy the crystal cages. Consequently, SDS electrophoresis of the dissolved crystals shows high molecular weight protein bands (Fig 3E). Since it is crucial that the protein extract doesńt become contaminated with tetrameric TTR, we decided to test other types of mesoporous crystals built with biomolecules – protein cross-linked crystals – with the expectation that they are less prone to surface adsorption of plasma proteins.

### 3.2. Thermolysin cross-linked crystals (TLN-CLC) separate TTR monomers from tetramers in vitro

TLN crystals grow in two main packing systems, hexagonal and tetragonal. These distributions can be achieved by modifying one component of the mixing solution: zinc (Hausrath and Matthews 2002). When grown in media that contain the ion (for example zinc chloride or zinc acetate), the crystals adopt a tetragonal packing system. In contrast, if the solution does not contain zinc, the packing will be hexagonal (Leite and Gales 2019). Tetragonal crystals are more uniform and have a solvent content of 66% (Fig. 4A). Hexagonal crystals are longer and rod-like and have a solvent content of 46% (Juers et al. 2018). Owing to the desired application of the crystals, the packing system of greatest interest is the tetragonal because it displays one-dimensional channels with appropriate dimensions along the *c*-crystallographic axis. Soakability predictions of crystal channels using *MAP_CHANNELS* (Juers and Ruffin 2014) and *LifeSoaks* (Pletzer-Zelgert et al. 2023) provided an overall bottleneck radius of 1.7 nm and a maximum pore radius of 2.0 nm. The surface of the TLN crystal channels show an overall hydrophobic environment and display only weak and localised charges. The target protein, TTR, in the native tetrameric form has a maximum dimension of 7.0 nm. Despite the lack of information about the structure and flexibility of monomeric TTR, the channel dimensions make tetragonal TLN crystals good candidates for extraction of the TTR monomeric species and size-exclusion of the tetramers (Fig. 4B). Furthermore, it is known that TTR dissociation exposes hydrophobic regions in the monomer surface (Kim et al. 2016) which probably drives diffusion from the bulk solution and potentiate pore entrapment; accordingly, the low charged surface of TLN channels generate an amenable chemical environment for uptake of TTR monomers.

**Fig. 4.**
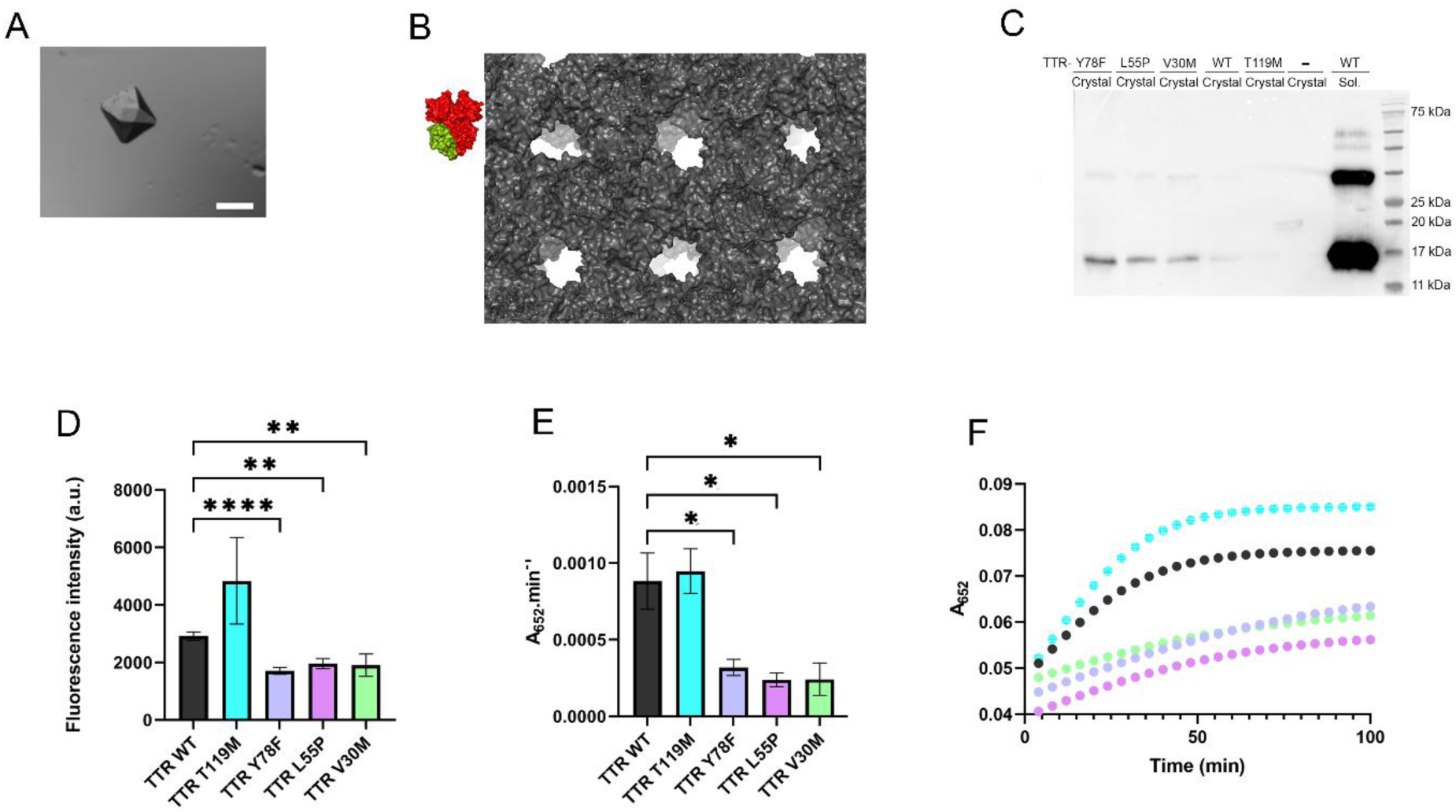
TLN-CLC efficiently extract TTR monomer from amyloidogenic TTR samples up to observable quantities. **A**. Optical image of TLN-CLC (scale bar 0.5 mm). **B**. Crystal structures of TTR with one of the monomers highlighted in green (PDB 1Y1D (Gales et al. 2005)) and of the TLN tetragonal framework (PDB 6N4Z (Harrison et al. 2019)). **C**. Western blotting analysis, with an antibody targeting human TTR, of the uptake by TLN-CLC crystals of TTR variants displaying a range of quaternary stability behaviours. **D** and **E**. Correlation between the crystal free void volumes after incubation with several TTR variants, estimated with fluorescein and cytochrome c, respectively. **F**. A colorimetric reaction was used to estimate cytochrome c trapped in the crystals (considered to be proportional to the reaction initial velocities). Examples of reaction progression curves used to estimate initial velocities are shown (color code of TTR variants in which the crystals were pre-incubated as in **E**. Three independent experiments were performed for each condition.

The packing of the grown crystals was confirmed by single crystal X-ray diffraction using an in-house equipment (Rigaku/Oxford Diffraction Gemini PX Ultra single-crystal X-ray diffractometer). Crystals were then cross-linked with glutaraldehyde, incubated with five recombinant TTR variants and then the protein uptake from each sample was analysed. The five TTR variants tested were three amyloidogenic mutants, TTR-Y78F, TTR-L55P and TTR-V30M, the TTR-WT and one variant with protective effect TTR-T119M (Almeida et al. 2000). Western blotting analysis using an antibody targeting human TTR shows that TTR uptake is higher for TTR-Y78F, TTR-L55P and TTR-V30M than for TTR-WT and TTR-T119M, and thus that it correlates with TTR quaternary instability (Fig 4C). The fact that TTR-T119M which is even more stable than TTR-WT is hardly detected gives a clear indication that the native TTR tetramer does not penetrate in the TLN-CLC framework.

In addition, we developed a cheaper alternative strategy to the use of anti-TTR antibodies to access the crystal extraction from pure recombinant TTR samples. Uptake of TTR monomers reduces the empty porous volume of the crystal that will be subsequently occupied by a secondary guest prone to optical sensing. Two secondary guests were tested. One is cytochrome c that has similar molecular dimensions to the TTR monomer and was already shown to almost fulfil the accessible void volume of MOF mesoporous crystals (Chen et al. 2012).If the same trend is observed with TLN-CLC crystals - almost complete occupation of the free void volume – it provides an indirect method to quantify monomeric TTR extraction. Cytochrome c catalyses a colorimetric reaction, and the initial velocity of the reaction catalysed by the crystals was used to access the crystal incorporation of this enzyme. The other probe tested is fluorescein, that should diffuse freely through the large pores of the TLN-CLC framework and equilibrate with the bulk solution concentration. Fluorescence intensity of this compound enables facile quantification of the uptake extent. The results of the assays that make use of the secondary probes are presented in Fig. 4 D and E. The secondary probes are inverted indicators of the monomeric TTR uptake: crystals pre-incubated with the amyloidogenic variants show lower uptake of the secondary probe (due to the occupation of the crystal void volume by monomeric TTR) when compared with the ones incubated with the stable TTR variants, which agrees with the western blot analysis (figure 3A). The assay is thus useful for facile and cheap sensing of monomers present in pure recombinant TTR samples. The results with the two secondary probes show a similar trend; there are not significant variations in the extraction of the three amyloidogenic variants tested. Extraction of TTR-T119M monomers looks slightly lower than the TTR-WT, as expected, due to the quaternary stabilizing role of this mutation.

### 3.3. TLN-CLC -based assay enables the evaluation the TTR drug candidates at nearly physiologic conditions

Small compounds, mostly binding in the TTR channel, are being developed to stabilize the protein native structure and therefore inhibit TTR amyloid formation. This therapeutic strategy is being persecuted for many years and, as a result, tafamidis became the first compound approved for the treatment of TTR amyloidosis by the European Commission and the FDA. Here, we evaluated the protective effect of two TTR stabilizers, Tafamidis and Acoramidis, at nearly physiologic conditions.

Tafamidis, or 2-(3,5-dichloro-phenyl)-benzoxazole-6-carboxylic acid, selectively binds TTR and kinetically stabilizes WT-TTR and mutant tetramers under denaturing and physiologic conditions, inhibiting amyloidogenesis (Bulawa et al. 2012). The authors overcame the difficulty to detect monomeric species under physiologic conditions indirectly, using subunit exchange experiments. A double-blind clinical trial demonstrated the efficacy of tafamidis in slowing the progression of TTR (Coelho et al. 2012). Acoramidis (AG10), or 3- [3-(3,5-dimethyl-1h-pyrazol-4-Yl)propoxy]-4-fluorobenzoic acid, is another promising drug that achieves near-complete stabilization of TTR in the plasma (Judge et al. 2019). It is currently in clinical trials for the treatment of TTR Amyloidosis (Fox et al. 2020).

The amyloidogenic proteins variants TTR-L55P and TTR-Y78F were used, and experiments were performed with drug / TTR molar ratios of 10/1, 1/1 and 0/1. The results are presented in Fig. 5. The two molecular probes (cytochrome c and fluorescein) used to estimate crystal void volumes yielded similar results (no statistically significant differences observed). In general, there was a significant reduction in the dissociation into monomers after addition of equimolar concentration of the drug; increasing 10x the concentration of the drug further increases the stability of the tetramer at physiological conditions. A drug / TTR molar ratio of 10/1, but not of 1/1, yielded dissociation rates equivalent to the TTR-WT. The assay is not sensible enough to detect significant differences between the action of tafamidis and acoramidis. Also, a consistent drug stabilizing effect was observed towards the two amyloidogenic variants.

**Fig. 5.**
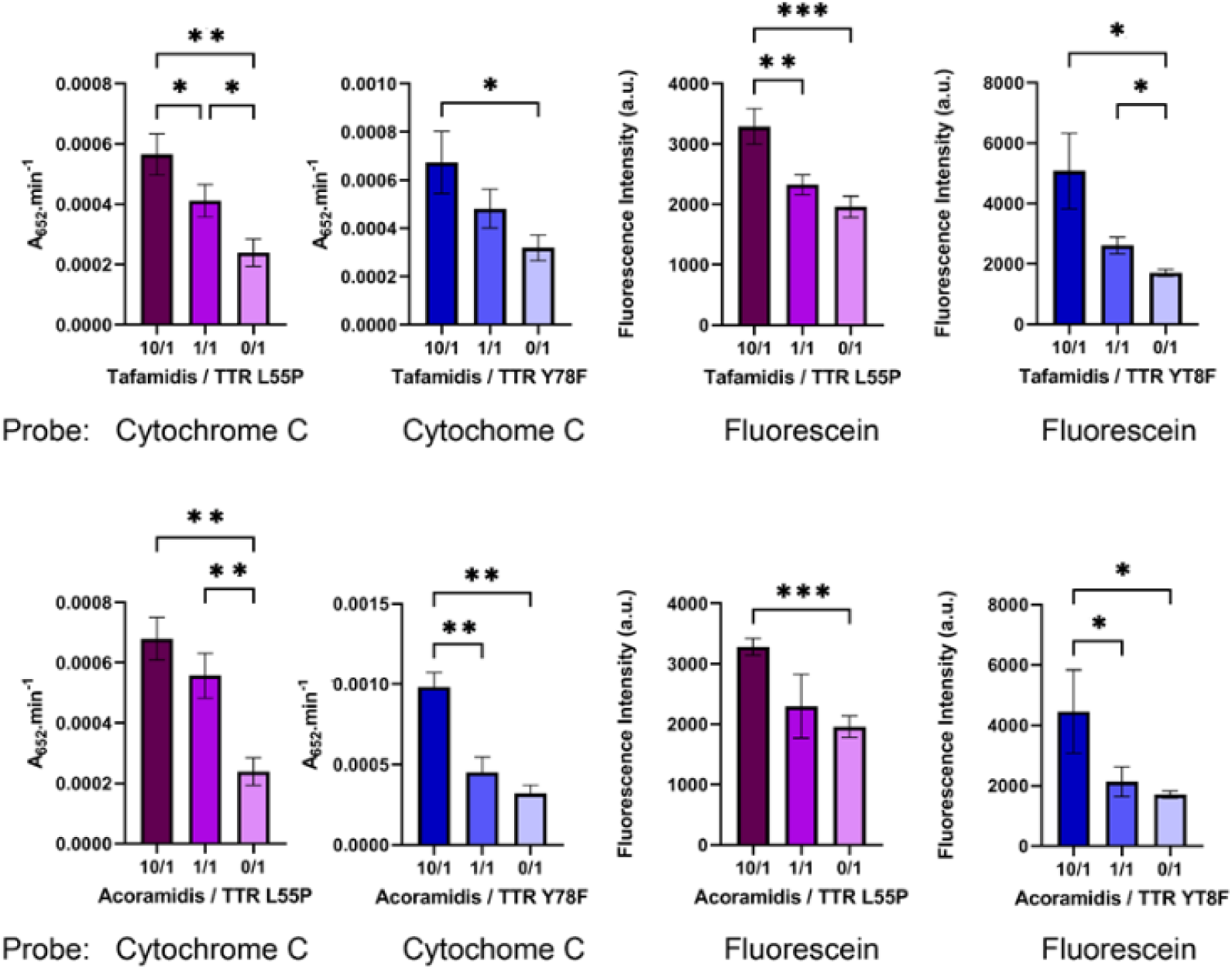
Application of TLN-CLC-based assay for evaluation of TTR stabilizers at nearly physiological conditions. Two drugs were tested, tafamidis (top) and acoramidis (bottom), with the amyloidogenic variants TTR-L55P and TTR-Y78F. Drug / TTR molar ratio: 10 /1, 1/1 and 0/1. TTR monomer uptake estimated indirectly, using secondary probes cytochrome c and fluorescein. Stability of the TTR variants is very sensitive do the drug / protein stoichiometry; the two drugs show a similar activity with both TTR variants. Results from three independent experiments for each condition.

Thus, we devised an in vitro assay for facile evaluation of the effect of compounds on TTR stability at physiologic pH. The assay outcomes the technical complexity of other indirect methods already in use, such as the determination of TTR subunit exchange rates (Nelson et al. 2021). Moreover, the assay is very sensitive to the drug / protein stoichiometry. It also reveals that stabilizing effect of the two compounds tested, tafamidis and acoramidis, in the amyloidogenic variants TTR-L55P and TTR-Y78F is very similar.

### 3.4. TLN-CLC - based assay reveals increased levels of TTR monomers in the plasma of TTR amyloidosis patients

Sensing monomeric TTR in plasma microsamples and proving the utility of the assay to discriminate between carriers of amyloidogenic TTR variants and TTR-WT constitute the main goal of this work. The TLN-CLC - based assay was tested with plasma samples of ten individuals, five carriers of TTR-V30M (# 53, 55, 56. 61 and 69) and five healthy controls (# 51, 52, 54. 57 and 58).

TLN-CLC crystals were incubated in 5 µl plasma samples for 3 h and the protein uptake analysed. We first checked by SDS analysis that unspecific adsorption of high molecular weight plasma proteins in the external surface of TLN-CLC crystals was negligible (not shown), providing a critical advantage towards the use of Tb-MOF crystals. This is crucial because the immunosensing assay that will be used subsequently for quantification of TTR collected from the crystals, does not efficiently discriminate between tetramers and monomers.

Identification of the target protein in the crystalś extracts was accomplished by anti-TTR Western blotting (Fig. 6A). Despite the variations observed in the monomeric TTR gel band intensities among the samples of TTR-V30M carriers, imaging analysis shows that they are significantly stronger than the ones of the controls. It should be noticed that semi-denaturing conditions were used, which enables the detection of the dimeric TTR band when a solution of TTR-WT is loaded (positive control). The TTR dimer is not detected in the crystal extracts which confirms that TTR surface adsorption is negligible.

**Fig. 6.**
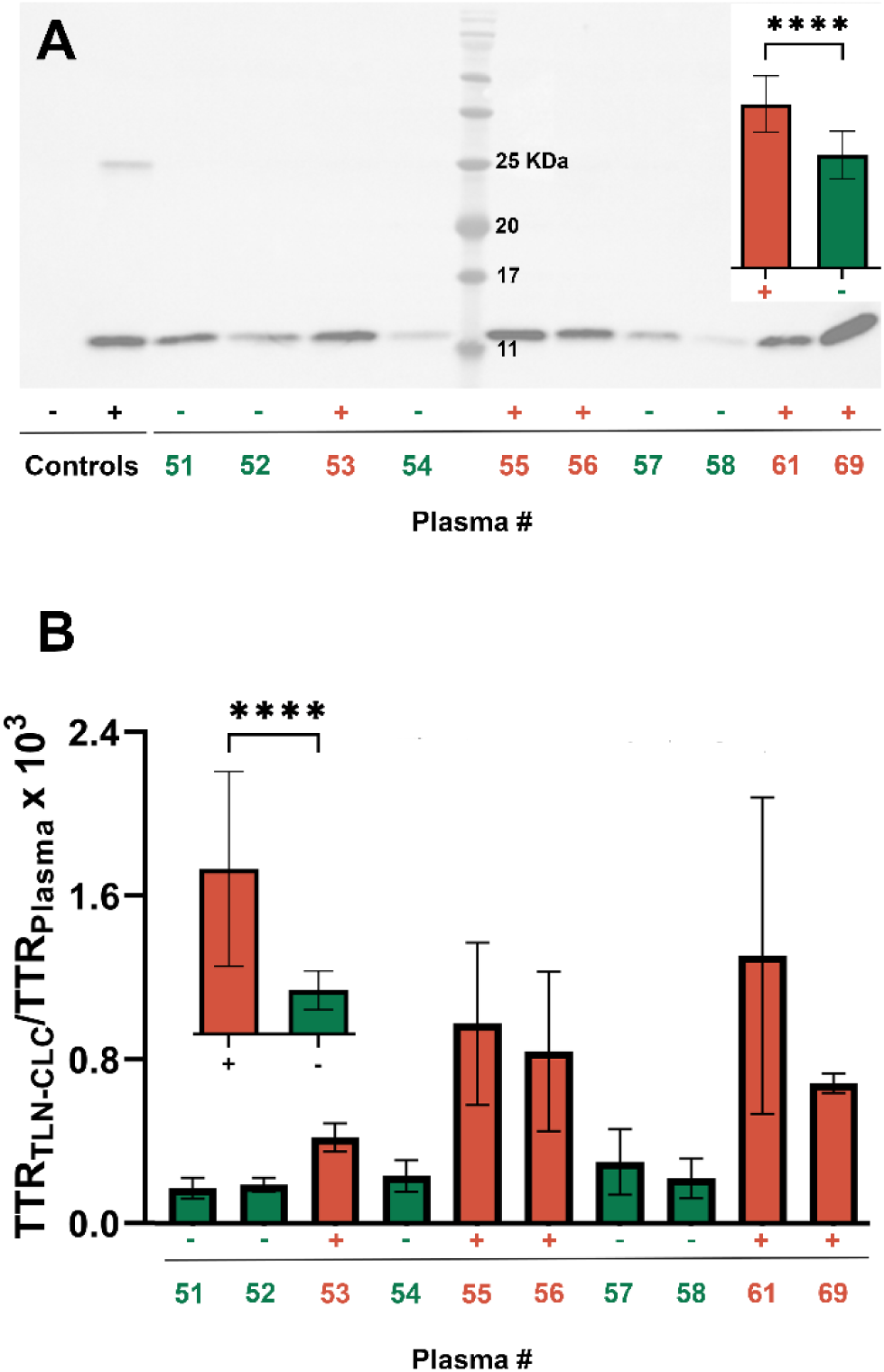
TLN-CLC - based assay reveals increased levels of TTR monomers in the plasma of TTR-V30M carriers. A. Representative anti-TTR Western blots of protein extracted by TLN-CLC crystals from plasma samples of TTR-V30M carriers (n= 5, +, red) and healthy individuals (n=5, -, green). Negative control: TLN-CLC crystal; Positive control: recombinant TTR-WT sample. Inset: Comparison between TTR band intensity between carriers and non-carriers of TTR-V30M determined by image analysis of the protein bands (four independent experiments for each plasma sample, 40 protein bands in total). B. Ratio of monomeric (extracted by the TLN-CLC)/ total TTR in plasma samples, quantified by ELISA. Inset: Comparison between the two groups of individuals, TTR-V30M carriers (red) and non-carriers (green). Three independent experiments were performed for each condition.

The monomeric TTR released from the TLN-CLC crystals, after incubation in plasma samples, was analysed by ELISA (Fig, 6B). In addition, the ELISA assay was also used to quantify the total concentration of TTR in plasma samples. It was observed that plasma TTR concentration of the TTR-V30M carriers is lower than the one of healthy controls, a feature already pointed for carriers of other amyloidogenic mutations such as TTR-V122I (Buxbaum et al. 2008). Moreover, it was also already noticed that age, gender and progression of the disease influence in a statistically significant manner the serum TTR levels (Buxbaum et al. 2008), which, together with the small sample size used here, may contribute to heterogeneity of the results. To exclude the effect of the variations in the levels of total plasma TTR, we found more reliable to use the monomeric TTR / total TTR ratio instead of the monomeric TTR alone.

The monomeric TTR / total TTR ratio was determined for the plasma samples of the 10 individuals (three independent experiments for each sample) and the results are presented in Fig. 6B. TLN-CLC crystals extract a fraction of 10^−4^ to 10^−3^ of the total TTR in the plasma samples. Comparison between the two groups of individuals, shows that the monomeric TTR/ total TTR ratio of the TTR-V30M carriers is higher than normal in a statistically significant manner. As expected, the difference towards the control group is bigger than the obtained by anti-TTR Western blotting of the monomers (insets of Fig. 6A and B) due to the normalization by the respective plasma concentration of total TTR. In line with our results, the presence of TTR oligomers circulating in the plasma of patients that are carriers of TTR-V30M in concentrations above healthy controls was already detected using peptide probes (Schonhoft et al. 2017). According to the established TTR amyloid cascade, the oligomers are formed by the self-assembly of monomers, not of native tetramers.

It is interesting to notice that no statistically significant variations were observed within the control group, an indication that the monomeric TTR / total TTR ratio eliminates gender and age caused variations, becoming a reliable indicator of disease progression. On the other hand, variations among the group of individuals carrying the TTR-V30M are much more pronounced, probably due to different stages in the disease progression within the group, a feature that could not be confirmed because the clinical state of the individuals carrying the amyloidogenic mutation was not disclosed.

## 4. Conclusion

Tetramer dissociation is the controlling step in TTR amyloidogenesis which prompted us to develop an assay to detect variations in the concentration of circulating monomeric TTR. The assay is technically simple to apply, demands a small volume of plasma sample (around 5 µl) and aims to sense a more suitable target (monomeric TTR) for early diagnosis and accurate evaluation of any therapeutic intervention than standard techniques, which analyse the deposition of amyloid aggregates. Moreover, the use of a fluid biomarker (circulating monomeric TTR) eliminates the currently adopted invasive biopsies modalities.

The assay was tested with two groups of five individuals each, one composed by carriers of TTR-V30M variant and the other by healthy controls. It was observed that TTR amyloidosis patients have significantly higher concentration of monomeric TTR than healthy controls. Moreover, determination of monomeric TTR / total TTR ratio was found to be more reliable than that of monomeric TTR alone. This is because total TTR concentration varies from individual to individual due to several factors, such as gender, age and the progression of the disease itself. The same is found in diagnostic of the Alzheimeŕs disease, in which the concentration of the more aggressive amyloid beta peptide 1-42 in the cerebrospinal fluid (CSF) is normalized by the basal production of amyloid beta peptide 1-40, that varies between individuals (Gales 2024).

Typical immunosensing assays rely on surface-capture of target molecules, but this constraint limits specificity towards the monomeric over the tetrameric TTR. We thus used instead mesoporous crystalline materials, which usually display excellent molecular sieving properties. While performing experiments with different materials we found much more challenging to work with plasma samples than with pure recombinant TTR samples, due to unspecific adsorption of plasma proteins to the external surface of the crystalline particles. In our test conditions, surface adsorption was much more severe in case of hybrid metal-organic materials than with protein-based materials. The later enabled us to develop the proposed assay. Implementation of the assay is facilitated by the availability from commercial sources of the protein selected to construct the porous framework, thermolysin, and by the ease to grow the crystals abundantly.

Quantification of monomeric TTR concentrations can be useful not only for aiding physicians in point-of-care diagnosis but also for following the response to emergent therapies focused on the administration of TTR stabilizers. In this context, circulating monomeric TTR should be the ideal response-to-treatment biomarker. Finally, we have also developed an optical assay that can be used to evaluate TTR drug candidates in vitro under nearly physiological conditions. This is an improvement towards standard methods that use critical experimental conditions, such as low pH, and to other demanding methods used to assess the residual monomeric TTR levels observed at physiological pH.

## Data Availability

All data produced in the present study are available upon reasonable request to the authors

## Acknowledgements

This work was supported by National funds through FCT—Fundação para a Ciência e a Tecnologia/Ministério da Ciência, Tecnologia e Ensino Superior in the framework of the projects “Institute for Research and Innovation in Health Sciences” (POCI-01-0145-FEDER-007274) and by European Union’s Horizon 2020 research and innovation programme under grant agreement No. 952334 (PhasAGE).

FCT is gratefully acknowledged for the PhD grant SFRH/BD/151016/2021 (to D.C.). Protein production and purification performed at the i3S Biochemical and Biophysical Technologies Platform. The mass spectrometry analysis done at the Proteomics i3S Scientific Platform with the assistance of Hugo Osório and the support of the Portuguese Mass Spectrometry Network (ROTEIRO/0028/2013; LISBOA-01-0145-FEDER-022125).

## Notes

### Competing Interest Statement

The authors have declared no competing interest.

### Funding Statement

This work was supported by National funds through FCT Fundacao para a Ciencia e a Tecnologia/Ministerio da Ciencia, Tecnologia e Ensino Superior in the framework of the projects Institute for Research and Innovation in Health Sciences (POCI-01-0145-FEDER-007274) and by European Union's Horizon 2020 research and innovation programme under grant agreement No. 952334 (PhasAGE).
FCT is gratefully acknowledged for the PhD grant SFRH/BD/151016/2021 (to D.C.). Protein production and purification performed at the i3S Biochemical and Biophysical Technologies Platform. The mass spectrometry analysis done at the Proteomics i3S Scientific Platform with the assistance of Hugo Osorio and the support of the Portuguese Mass Spectrometry Network (ROTEIRO/0028/2013; LISBOA-01-0145-FEDER-022125).

### Author Declarations

human blood samples from individuals who requested to perform blood collection with predictive character at the Center for Predictive and Preventive Genetics (CGPP) of the IBMC to evaluate the condition of the pre-symptomatic V30M mutation was submitted by the group Molecular Neurobiology (i3S-Instituto de Investigacao e Inovacao em Saude) and approved by the Ethical Committee of the University of Porto (Report n 36/CEUP/2017).

## References

Abe, S., Tabe, H., Ijiri, H., Yamashita, K., Hirata, K., Atsumi, K., Shimoi, T., Akai, M., Mori, H., Kitagawa, S., Ueno, T., 2017. Crystal Engineering of Self-Assembled Porous Protein Materials in Living Cells. ACS Nano 11(3), 2410–2419.

Adams, D., Suhr, O.B., Hund, E., Obici, L., Tournev, I., Campistol, J.M., Slama, M.S., Hazenberg, B.P., Coelho, T., 2016. First European consensus for diagnosis, management, and treatment of transthyretin familial amyloid polyneuropathy. Curr Opin Neurol 29 Suppl 1(Suppl 1), S14–26.

Afonso, R.V., Durão, J., Mendes, A., Damas, A.M., Gales, L., 2010. Dipeptide crystals as excellent permselective materials: Sequential exclusion of argon, nitrogen, and oxygen. Angewandte Chemie - International Edition 49(17), 3034–3036.

Almeida, M.R., Alves, I.L., Terazaki, H., Ando, Y., Saraiva, M.J., 2000. Comparative studies of two transthyretin variants with protective effects on familial amyloidotic polyneuropathy: TTR R104H and TTR T119M. Biochem Biophys Res Commun 270(3), 1024–1028.

Almeida, M.R., Gales, L., Damas, A.M., Cardoso, I., Saraiva, M.J., 2005. Small transthyretin (TTR) ligands as possible therapeutic agents in TTR amyloidoses. Curr Drug Targets CNS Neurol Disord 4(5), 587–596.

Benson, M.D., Waddington-Cruz, M., Berk, J.L., Polydefkis, M., Dyck, P.J., Wang, A.K., Planté-Bordeneuve, V., Barroso, F.A., Merlini, G., Obici, L., Scheinberg, M., Brannagan, T.H., Litchy, W.J., Whelan, C., Drachman, B.M., Adams, D., Heitner, S.B., Conceição, I., Schmidt, H.H., Vita, G., Campistol, J.M., Gamez, J., Gorevic, P.D., Gane, E., Shah, A.M., Solomon, S.D., Monia, B.P., Hughes, S.G., Jesse Kwoh, T., McEvoy, B.W., Jung, S.W., Baker, B.F., Ackermann, E.J., Gertz, M.A., Coelho, T., 2018. Inotersen treatment for patients with Hereditary transthyretin amyloidosis. New England Journal of Medicine 379(1), 22–31.

Bonifácio, M.J., Sakaki, Y., Saraiva, M.J., 1996. ’In vitro’ amyloid fibril formation from transthyretin: The influence of ions and the amyloidogenicity of TTR variants. Biochimica et Biophysica Acta - Molecular Basis of Disease 1316(1), 35–42.

Bulawa, C.E., Connelly, S., DeVit, M., Wang, L., Weigel, C., Fleming, J.A., Packman, J., Powers, E.T., Wiseman, R.L., Foss, T.R., Wilson, I.A., Kelly, J.W., Labaudinière, R., 2012. Tafamidis, a potent and selective transthyretin kinetic stabilizer that inhibits the amyloid cascade. Proceedings of the National Academy of Sciences of the United States of America 109(24), 9629–9634.

Buxbaum, J., Koziol, J., Connors, L.H., 2008. Serum transthyretin levels in senile systemic amyloidosis: effects of age, gender and ethnicity. Amyloid 15(4), 255–261.

Chen, Y., Lykourinou, V., Vetromile, C., Hoang, T., Ming, L.J., Larsen, R.W., Ma, S., 2012. How can proteins enter the interior of a MOF? investigation of cytochrome c translocation into a MOF consisting of mesoporous cages with microporous windows. Journal of the American Chemical Society 134(32), 13188–13191.

Coelho, T., Maia, L.F., Martins da Silva, A., Waddington Cruz, M., Planté-Bordeneuve, V., Lozeron, P., Suhr, O.B., Campistol, J.M., Conceição, I.M., Schmidt, H.H., Trigo, P., Kelly, J.W., Labaudinière, R., Chan, J., Packman, J., Wilson, A., Grogan, D.R., 2012. Tafamidis for transthyretin familial amyloid polyneuropathy: a randomized, controlled trial. Neurology 79(8), 785–792.

Comotti, A., Bracco, S., Distefano, G., Sozzani, P., 2009. Methane, carbon dioxide and hydrogen storage in nanoporous dipeptide-based materials. Chemical Communications(3), 284–286.

Cvetkovic, A., Picioreanu, C., Straathof, A.J.J., Krishna, R., Van Der Wielen, L.A.M., 2005. Relation between pore sizes of protein crystals and anisotropic solute diffusivities. Journal of the American Chemical Society 127(3), 875–879.

Dong, X., Zhao, G., Li, X., Fang, J., Miao, J., Wei, Q., Cao, W., 2020. Electrochemiluminescence immunosensor of “signal-off” for beta-amyloid detection based on dual metal-organic frameworks. Talanta 208, 120376.

Durão, J., Gales, L., 2013. Guest diffusion in dipeptide crystals. CrystEngComm 15(8), 1532–1535.

Fox, J.C., Hellawell, J.L., Rao, S., O’Reilly, T., Lumpkin, R., Jernelius, J., Gretler, D., Sinha, U., 2020. First-in-Human Study of AG10, a Novel, Oral, Specific, Selective, and Potent Transthyretin Stabilizer for the Treatment of Transthyretin Amyloidosis: A Phase 1 Safety, Tolerability, Pharmacokinetic, and Pharmacodynamic Study in Healthy Adult Volunteers. Clinical Pharmacology in Drug Development 9(1), 115–129.

Furuya, H., Saraiva, M.J.M., Gawinowicz, M.A., Alves, I.L., Costa, P.P., Sasaki, H., Goto, I., Sakaki, Y., 1991. Production of recombinant human transthyretin with biological activities toward the understanding of the molecular basis of familial amyloidotic polyneuropathy (FAP). Biochemistry 30(9), 2415–2421.

Gales, L., 2019. Tegsedi (Inotersen): An Antisense Oligonucleotide Approved for the Treatment of Adult Patients with Hereditary Transthyretin Amyloidosis. Pharmaceuticals (Basel) 12(2).

Gales, L., 2024. Detection and clearance in Alzheimer’s disease: leading with illusive chemical, structural and morphological features of the targets. Neural Regeneration Research 19(3), 497–498.

Gales, L., Macedo-Ribeiro, S., Arsequell, G., Valencia, G., Saraiva, M.J., Damas, A.M., 2005. Human transthyretin in complex with iododiflunisal: Structural features associated with a potent amyloid inhibitor. Biochemical Journal 388(2), 615–621.

Görbitz, C.H., 2001. Nanotube formation by hydrophobic dipeptides. Chemistry - A European Journal 7(23), 5153–5159.

Gu, C., Liu, Y., Hu, B., Liu, Y., Zhou, N., Xia, L., Zhang, Z., 2020. Multicomponent nanohybrids of nickel/ferric oxides and nickel cobaltate spinel derived from the MOF-on-MOF nanostructure as efficient scaffolds for sensitively determining insulin. Anal. Chim. Acta 1110, 44–55.

Harrison, K., Wu, Z., Juers, D.H., 2019. A comparison of gas stream cooling and plunge cooling of macromolecular crystals. Journal of Applied Crystallography 52(5), 1222–1232.

Hashimoto, T., Ye, Y., Matsuno, A., Ohnishi, Y., Kitamura, A., Kinjo, M., Abe, S., Ueno, T., Yao, M., Ogawa, T., Matsui, T., Tanaka, Y., 2019. Encapsulation of biomacromolecules by soaking and co-crystallization into porous protein crystals of hemocyanin. Biochemical and Biophysical Research Communications 509(2), 577–584.

Hausrath, A.C., Matthews, B.W., 2002. Thermolysin in the absence of substrate has an open conformation. Acta Crystallographica Section D 58(6 Part 2), 1002–1007.

Judge, D.P., Heitner, S.B., Falk, R.H., Maurer, M.S., Shah, S.J., Witteles, R.M., Grogan, M., Selby, V.N., Jacoby, D., Hanna, M., Nativi-Nicolau, J., Patel, J., Rao, S., Sinha, U., Turtle, C.W., Fox, J.C., 2019. Transthyretin Stabilization by AG10 in Symptomatic Transthyretin Amyloid Cardiomyopathy. Journal of the American College of Cardiology 74(3), 285–295.

Juers, D.H., Farley, C.A., Saxby, C.P., Cotter, R.A., Cahn, J.K.B., Holton-Burke, R.C., Harrison, K., Wu, Z., 2018. The impact of cryosolution thermal contraction on proteins and protein crystals: volumes, conformation and order. Acta Crystallographica Section D 74(9), 922–938.

Juers, D.H., Ruffin, J., 2014. MAP-CHANNELS: A computation tool to aid in the visualization and characterization of solvent channels in macromolecular crystals. Journal of Applied Crystallography 47(6), 2105–2108.

Kim, J.H., Oroz, J., Zweckstetter, M., 2016. Structure of Monomeric Transthyretin Carrying the Clinically Important T119M Mutation. Angewandte Chemie International Edition 55(52), 16168–16171.

Kowalski, A.E., Johnson, L.B., Dierl, H.K., Park, S., Huber, T.R., Snow, C.D., 2019. Porous protein crystals as scaffolds for enzyme immobilization. Biomaterials Science 7(5), 1898–1904.

Lai, Y.T., Reading, E., Hura, G.L., Tsai, K.L., Laganowsky, A., Asturias, F.J., Tainer, J.A., Robinson, C.V., Yeates, T.O., 2014. Structure of a designed protein cage that self-assembles into a highly porous cube. Nat Chem 6(12), 1065–1071.

Leite, J.P., Figueira, F., Mendes, R.F., Almeida Paz, F.A., Gales, L., 2023. Metal–Organic Frameworks as Sensors for Human Amyloid Diseases. ACS Sensors 8(3), 1033–1053.

Leite, J.P., Gales, L., 2019. Alzheimer’s Aβ1-40 peptide degradation by thermolysin: evidence of inhibition by a C-terminal Aβ product. FEBS Letters 593(1), 128–137.

Leite, J.P., Rodrigues, D., Ferreira, S., Figueira, F., Almeida Paz, F.A., Gales, L., 2019. Mesoporous Metal–Organic Frameworks as Effective Nucleating Agents in Protein Crystallography. Crystal Growth & Design 19(3), 1610–1615.

Mendes, R.F., Figueira, F., Leite, J.P., Gales, L., Almeida Paz, F.A., 2020. Metal-organic frameworks: a future toolbox for biomedicine? Chem Soc Rev 49(24), 9121–9153.

Nelson, L.T., Paxman, R.J., Xu, J., Webb, B., Powers, E.T., Kelly, J.W., 2021. Blinded potency comparison of transthyretin kinetic stabilisers by subunit exchange in human plasma. Amyloid 28(1), 24–29.

Parakra, R.D., Kleffmann, T., Jameson, G.N.L., Ledgerwood, E.C., 2018. The proportion of Met80-sulfoxide dictates peroxidase activity of human cytochrome c. Dalton Transactions 47(27), 9128–9135.

Park, Y.K., Sang, B.C., Kim, H., Kim, K., Won, B.H., Choi, K., Choi, J.S., Ahn, W.S., Won, N., Kim, S., Dong, H.J., Choi, S.H., Kim, G.H., Cha, S.S., Young, H.J., Jin, K.Y., Kim, J., 2007. Crystal structure and guest uptake of a mesoporous metal-organic framework containing cages of 3.9 and 4.7 nm in diameter. Angewandte Chemie - International Edition 46(43), 8230–8233.

Pletzer-Zelgert, J., Ehrt, C., Fender, I., Griewel, A., Flachsenberg, F., Klebe, G., Rarey, M., 2023. LifeSoaks: A tool for analyzing solvent channels in protein crystals and obstacles for soaking experiments. Acta Crystallographica Section D: Structural Biology 79, 837–856.

Redondo, C., Damas, A.M., Olofsson, A., Lundgren, E., Saraiva, M.J.M., 2000. Search for intermediate structures in transthyretin fibrillogenesis: Soluble tetrameric Tyr78Phe TTR expresses a specific epitope present only in amyloid fibrils. Journal of Molecular Biology 304(3), 461–470.

Schonhoft, J.D., Monteiro, C., Plate, L., Eisele, Y.S., Kelly, J.M., Boland, D., Parker, C.G., Cravatt, B.F., Teruya, S., Helmke, S., Maurer, M., Berk, J., Sekijima, Y., Novais, M., Coelho, T., Powers, E.T., Kelly, J.W., 2017. Peptide probes detect misfolded transthyretin oligomers in plasma of hereditary amyloidosis patients. Science Translational Medicine 9(407).

Sheta, S.M., El-Sheikh, S.M., Abd-Elzaher, M.M., 2019. A novel optical approach for determination of prolactin based on Pr-MOF nanofibers. Anal. Bioanal. Chem. 411(7), 1339–1349.

Song, X., Shao, X., Dai, L., Fan, D., Ren, X., Sun, X., Luo, C., Wei, Q., 2020. Triple Amplification of 3,4,9,10-Perylenetetracarboxylic Acid by Co(2+)-Based Metal-Organic Frameworks and Silver-Cysteine and Its Potential Application for Ultrasensitive Assay of Procalcitonin. ACS Appl. Mater. Interfaces 12(8), 9098–9106.

Tajahmadi, S., Molavi, H., Ahmadijokani, F., Shamloo, A., Shojaei, A., Sharifzadeh, M., Rezakazemi, M., Fatehizadeh, A., Aminabhavi, T.M., Arjmand, M., 2023. Metal-organic frameworks: A promising option for the diagnosis and treatment of Alzheimer’s disease. Journal of Controlled Release 353, 1–29.

Vilenchik, L.Z., Griffith, J.P., St. Clair, N., Navia, M.A., Margolin, A.L., 1998. Protein Crystals as Novel Microporous Materials. Journal of the American Chemical Society 120(18), 4290–4294.

Wang, Z., Hu, X., Sun, N., Deng, C., 2019. Aptamer-functionalized magnetic metal organic framework as nanoprobe for biomarkers in human serum. Anal. Chim. Acta 1087, 69–75.

Wu, Q., Tan, R., Mi, X., Tu, Y., 2020. Electrochemiluminescent aptamer-sensor for alpha synuclein oligomer based on a metal-organic framework. Analyst 145(6), 2159–2167.

Zhou, Y., Li, C., Li, X., Zhu, X., Ye, B., Xu, M., 2018. A sensitive aptasensor for the detection of β-amyloid oligomers based on metal–organic frameworks as electrochemical signal probes. Anal. Methods 10(36), 4430–4437.

